# Integrative transcriptome analysis of malignant pleural mesothelioma reveals a clinically relevant immune-based classification

**DOI:** 10.1101/2020.08.14.20174789

**Authors:** Ania Alay, David Cordero, Sara Hijazo-Pechero, Elisabet Aliagas, Adriana Lopez-Doriga, Raúl Marín, Ramón Palmero, Roger Llatjós, Ignacio Escobar, Ricard Ramos, Susana Padrones, Víctor Moreno, Ernest Nadal, Xavier Solé

## Abstract

**Background:** Malignant pleural mesothelioma (MPM) is a rare and aggressive neoplasia affecting the lung mesothelium. Immune checkpoint inhibitors (ICI) in MPM have not been extremely successful, likely due to poor identification of suitable candidate patients for the therapy. We aimed to identify cellular immune fractions associated with clinical outcome and classify MPM patients based on their immune contexture. For each defined group, we sought for molecular specificities that could help further define our MPM classification at the genomic and transcriptomic level, as well as identify differential therapeutic strategies based on transcriptional signatures predictive of drug response.

**Methods:** The abundance of 20 immune cell fractions in 516 MPM samples from 7 gene expression datasets was inferred using Gene Set Variation Analysis. Identification of clinically relevant fractions was performed with Cox Proportional-Hazards Models adjusted for age, stage, sex, and tumor histology. Immune-based groups were identified using unsupervised classification.

**Results:** T-Helper 2 (T_H2_) and cytotoxic T (T_C_) cells were found to be consistently associated with overall survival. Three immune clusters (IG) were subsequently defined based on T_H2_ and T_C_ immune infiltration levels: IG1 (54.5%) was characterized by high T_H2_ and low T_C_ levels, IG2 (37%) had either low or high levels of both fractions, and IG3 (8.5%) was defined by low T_H2_ and high T_C_ levels. IG1 and IG3 groups were associated with worse and better overall survival, respectively. The three groups showed differential molecular profiles, with IG1 enriched for *CDKN2A* and IFN-related genes deletions. At the transcriptional level, IG1 samples showed upregulation of proliferation signatures, while IG3 samples presented upregulation of immune and inflammation-related pathways. Finally, the integration of gene expression with functional signatures of drug response showed that IG3 patients might be more likely to respond to ICI, while IG1 patients could be more sensitive to PARP inhibitors.

**Conclusions:** This study identifies a novel immune-based signature with potential clinical relevance based on T_H2_ and T_C_ levels, unveiling a fraction of MPM patients with better prognosis and who might benefit from immune-based therapies. Genomic and transcriptomic specificities of the different groups could be used to tailor potential therapies in the future.

## BACKGROUND

Malignant pleural mesothelioma (MPM) is a rare and highly aggressive neoplasia arising in the pleural cavity, and its incidence is increasing worldwide. MPM tumors are classified into three distinct histological subtypes: epithelioid, biphasic, and sarcomatoid. Median overall survival in MPM patients is approximately 12 months, although patients with epithelioid tumors usually have better prognosis than individuals with biphasic or sarcomatoid MPM.[1] Furthermore, most patients are diagnosed in advanced stages, which still aggravates the burden caused by the disease. The use of surgery in MPM patients is very limited, and current standard of care treatment for MPM patients is chemotherapy with platinum combined with pemetrexed, which has been shown to improve overall survival and quality of life.[2] However, benefits from chemotherapy are generally modest and prognosis remains dismal with 5-year survival rates lower than 5%.[3] Furthermore, there is no standard second line treatment when tumor progresses to front-line chemotherapy.

Immunotherapy based on monoclonal antibodies against programmed cell death protein 1 (PD-1) and programmed death ligand 1 (PD-L1) have been tested in clinical trials in MPM. Several nonrandomized phase I/II trials testing single-agent immune checkpoint inhibitors showed antitumor activity (9-29%) and median progression-free survival ranging from 2.8 to 6.2 months.[4] Preliminary results from phase II clinical trials combining in second line inhibitors of cytotoxic T-lymphocyte-associated antigen 4 (CTLA4) plus anti-PD1/PD-L1, such as ipilimumab and nivolumab or tremelimumab and durvalumab, showed promising efficacy results and also significant toxicity.[5] In those clinical trials, PD-L1 expression had limited value to predict benefit from immune checkpoint inhibitors. Moreover, tumor mutational burden (TMB) which is generally low in MPM[6] is not likely to become a predictive marker in this tumor. Therefore, it is essential to further study the role of the different components of the immune system in the evolution and prognosis of MPM, as well as the identification of additional biomarkers of immunotherapy response beyond PD-L1 immuno-histochemical expression.

In this study, our aim is to capture the landscape of the immune microenvironment in a large collection of 516 MPM gene expression profiles comprised in seven different publicly available datasets.[6-12] Once the immune contexture is determined for every tumor, we will identify those immune cell fractions with potential prognostic ability and classify MPM samples based on these previously identified clinically-relevant immune cell types. We will subsequently use the defined MPM groups to identify genomic and transcriptomic group specificities and evaluate functional signatures of drug response to suggest possible therapeutic strategies.

## METHODS

### Data collection and processing

A systematic search to identify publicly available datasets was performed in January 2018 using the query “pleural mesothelioma” and filtering to keep only human samples and expression profiling experiments in multiple open access repositories. We also filtered datasets with less than 30 samples and kept experiments that covered most of the transcriptome. Raw data were downloaded whenever possible and subsequently processed accordingly as summarized hereafter: for whole-exome sequencing data, Broad’s Genome Analysis Toolkit best practices were followed,[13] RNA-sequencing data was processed according to an in-house pipeline based on ENCODE best practices, and for expression arrays, robust multiarray average algorithm[14] was used to normalize the data. A detailed description of data identification and data processing is available in extended methods.

### Association of immune fractions with overall survival

The immune infiltrate composition of 20 different cell types was deconvoluted from the obtained gene expression profiles using Gene Set Variation Analysis (GSVA) method.[15] Gene signatures of the different immune fractions were obtained from a previous study[16] in which signatures for a total of 28 different immune cell types were generated, including innate immune cells (e.g., dendritic cells, eosinophils, mast cells, macrophages, natural killer cells, neutrophils) and adaptive immune cells (e.g., B cells, T cells). Since GSVA can only deconvolute multi-gene signatures, we replaced two single-gene signatures (plasmacytoid dendritic cells and regulatory T cells) with their corresponding multi-gene signatures from another study.[17] Additionally, when more specific categories were available for a cellular fraction (e.g. NK CD56dim and NK CD56bright), general categories (e.g. NK cells) were removed to avoid signature redundancy.

Once relative abundances for each immune fraction and sample where obtained, Cox proportional-hazards models adjusted for sex, stage, age, and histology were fitted using the largest dataset with available survival information as the discovery dataset,[6] and setting the optimal cut point that maximized survival differences for each immune fraction using R packages *survival* (v.3.1.7) and *survMisc* (v.0.5.5). Using the GSVA cut points defined in the discovery dataset, Cox proportional-hazards models adjusted for age, stage, sex, and histology were fitted in two additional validation datasets.[7,10] Fractions associated to clinical outcome were determined as those significant (p < 0.05) after FDR adjustment in the discovery dataset and at least one validation dataset, and with a consistent hazard ratio across the three datasets.

### Group characterization

#### Clinicopathological variables

Association with clinicopathological variables (age, sex, stage, tumor histology, asbestos exposure, and neoadjuvant therapy) was done using *compareGroups* package for R (v.4.2.0). For categorical variables, a Chi-squared test was performed, and continuous variables were tested using Kruskal-Wallis test.

#### Immune checkpoints and T cell exhaustion status

To assess immune checkpoints status and T cell exhaustion status in the different immune groups, multiple sets of genes were obtained from previous studies, including lists of immune checkpoint activators, immune checkpoint inhibitors,[18] and T cell exhaustion and effector markers.[19] For each gene, we performed a linear model adjusted for sex, age, stage, and histology in each dataset, where the immune group was codified as a numeric variable and the gene expression was the dependent variable. Datasets with less than three samples in any immune group were discarded. The weighted Z method from the *survcomp* package (v.1.36.0) was used to combine the p-values obtained for the different datasets. Bonferroni adjusted p-values lower than 0.05 were considered as significant. β values were combined using the weighted mean.

#### Genomic alterations

Both Bueno et al. and Hmeljak et al. datasets had somatic variant data available to evaluate the mutational burden. The total number of variants per sample was calculated excluding non-exonic and synonymous mutations. Using somatic single nucleotide variants, mutational signatures were inferred using the R package *deconstructSigs* (v.1.8.0). Neoepitope binding affinity was also derived from somatic single nucleotide variants. More details on these analyses are available in extended methods.

Copy number alteration burden was evaluated in the samples from the Hmeljak et al. dataset. Fisher tests were computed to assess the differences between immune groups in terms of number of altered/lost/gained genes. R package *copynumber* (v.1.24.0) was used to plot the genome overview of altered samples.

#### Transcriptomic analysis

The association of the expression levels of individual genes with the immune groups was done as described in the immune checkpoint evaluation section. For pathway analysis, a pre-ranked Gene Set Enrichment Analysis (GSEA) was run with genes ranked using the product of the slope and the negative logio(p-value) from the linear model used to assess transcriptomic status at the single-gene level. Gene sets tested included hallmarks and canonical pathways (KEGG, Biocarta, Reactome) from MSigDB version 6.1.[20]

#### Assessment of response to therapy

Transcriptional signatures of sensitivity or resistance to specific treatments were used to compute GSVA scores for each sample. The tested signatures were: pemetrexed resistance,[21] cisplatin resistance,[22] PARP inhibitors sensitivity,[23] and palbociclib resistance,[24] as well as multiple signatures predictive of benefit to immunotherapy found after a systematic search literature. A total of seven previously published studies with publicly available transcriptomic signatures were found.[25-31] Signatures with less than 10 genes were filtered out and linear models were fitted to obtain the slope of the signature correlation with the immune groups. Analysis of variance was used to test for significance of the models adjusted for histology and dataset.

## RESULTS

### Profiling of immune fractions in a large cohort of MPM tumors leads to clinically relevant immune cell populations

In order to identify clinically relevant immune-cell fractions in MPM tumors, we collected data from seven MPM gene expression datasets that fulfilled our selection criteria.[6-12] Overall, we collected 516 MPM gene expression profiles, with available survival information and covariates for 300 patients (58%). A summary of the clinicopathological characteristics of the samples in each dataset is available in Suppl. Table S1.

For each sample included in the analysis, we quantified the abundance of 20 immune-cell populations by applying GSVA[15] on a previously established set of immune-specific gene signatures (Suppl. Table S2).[16,17] Thus, for each sample and immune cellular fraction, we obtained a score between [-1, +1], with extreme values close to 1 or −1 indicating a strong presence or absence of a fraction in a specific sample, respectively. Similarly, GSVA scores close to 0 correspond to an unbiased distribution (i.e., centered or randomly distributed) of the genes across the whole gene expression profile.

Once the abundance of all immune fractions was quantified in the full cohort of samples, we aimed to assess whether there existed any individual fractions associated with overall patient survival using Cox proportional-hazards regression models adjusted for potential confounders (i.e., age, sex, tumor stage, histology, and dataset). Thus, for each cell type we identified the cut point that maximized the survival differences between high vs. low using the largest available dataset (Bueno et al.[6]). The cut points obtained in the discovery dataset were further validated in the two additional cohorts with available survival information (Hmeljak et al.[7] and Bott el al.[10]). Out of the 20 studied immune cell types, T helper 2 (T_H2_) and cytotoxic T cells (T_C_) showed a consistent association with patient survival across datasets based on our criteria (Cox p < 0.05 in at least 2 of the three datasets, and the remaining dataset showing a consistent trend, albeit not necessarily significant). In our analysis, higher levels of T_H2_ cells (GSVA score > −0.16) were associated with worse outcome (HR_Bueno_=2.1, p=0.00015; HR_Hmeijak_=2.6, p=0.0021; HR_Bott_=2.3, p=0.066; HR_Combined_=2.1, p=7.610^-7^) (Fig. 1A), while higher levels of T_C_ (GSVA score > 0.40) were associated with better prognosis (HR_Bueno_=0.57, p=0.0091; HR_Hmeijak_=0.73, p=0.39; HR_Bott_=0.29, p=0.016; HR_Combined_=0.59, p=0.00094) (Fig. 1B). Survival analysis results for the complete set of immune fractions can be found in Suppl. Fig. S1. When individuals were stratified in four groups based on the combination of the abundance (i.e., low vs. high) of both T_H2_ and T_C_ cells, MPM patients with low abundance of T_H2_ and high T_C_ infiltration consistently showed the best overall survival when compared to patients with high levels of T_H2_ and low levels of T_C_ (HR_Bueno_=0.34, p=0.00022; HR_Hmeijak_=0.20, p=0.0080; HR_Bott_=0.12, p=0.020; HR_Combined_=0.31, p=2.710^-7^) (Fig. 1C). Additionally, patients with either both high or low abundance levels of both cell types showed intermediate prognosis, suggesting some type of compensatory effect between these two immune cell types. Therefore, we decided to merge these two groups into a single category to create a final classification of three immune-based groups: T_H2_High-T_C_Low (IG1, worse prognosis, 54.5% of the patients), T_H2_High-T_C_High or T_H2_Low-T_C_Low (IG2, intermediate prognosis, 37% of the patients, HR_Combined_=0.52), T_H2_Low-T_C_High (IG3, better prognosis, 8.5% of the patients, HR_Combined_=0.32) (Fig. 1D).

**Figure 1.**
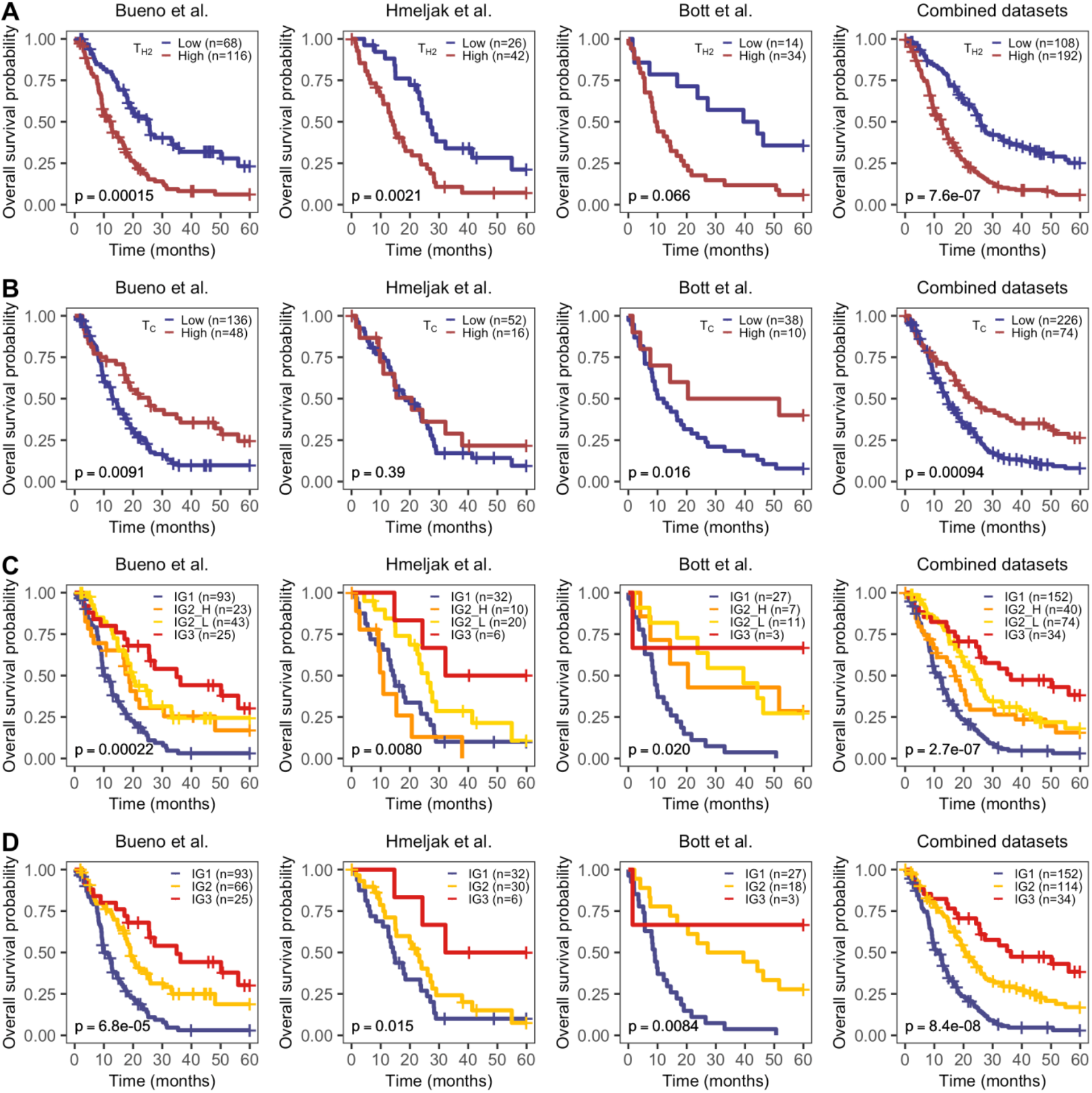
Overall survival by significant immune fractions. Kaplan-Meier curves of T-helper 2 cells (A), cytotoxic T cells (B), the combination of both fractions in 4 groups (C), and in 3 groups (D). Hazard ratios (HR) and p-values come from a Cox proportional-hazards model adjusted for age, sex, stage, and histology.

Once the immune-based MPM groups were defined, we aimed to identify any potential differences in the remaining immune fractions among them. Groups IG1 and IG2 comprised a mixture of samples with both strongly and less immune-enriched tumors, while on average group IG3 showed significant higher abundancefor 13 of the 20 immune cell fractions (Fig. 2A, Suppl. Fig. S2). These results suggest that the combination of these two immune fractions interplays with the remaining immune cell types and contributes to identify a subset of highly immune-infiltrated MPM tumors with improved prognosis.

**Figure 2.**
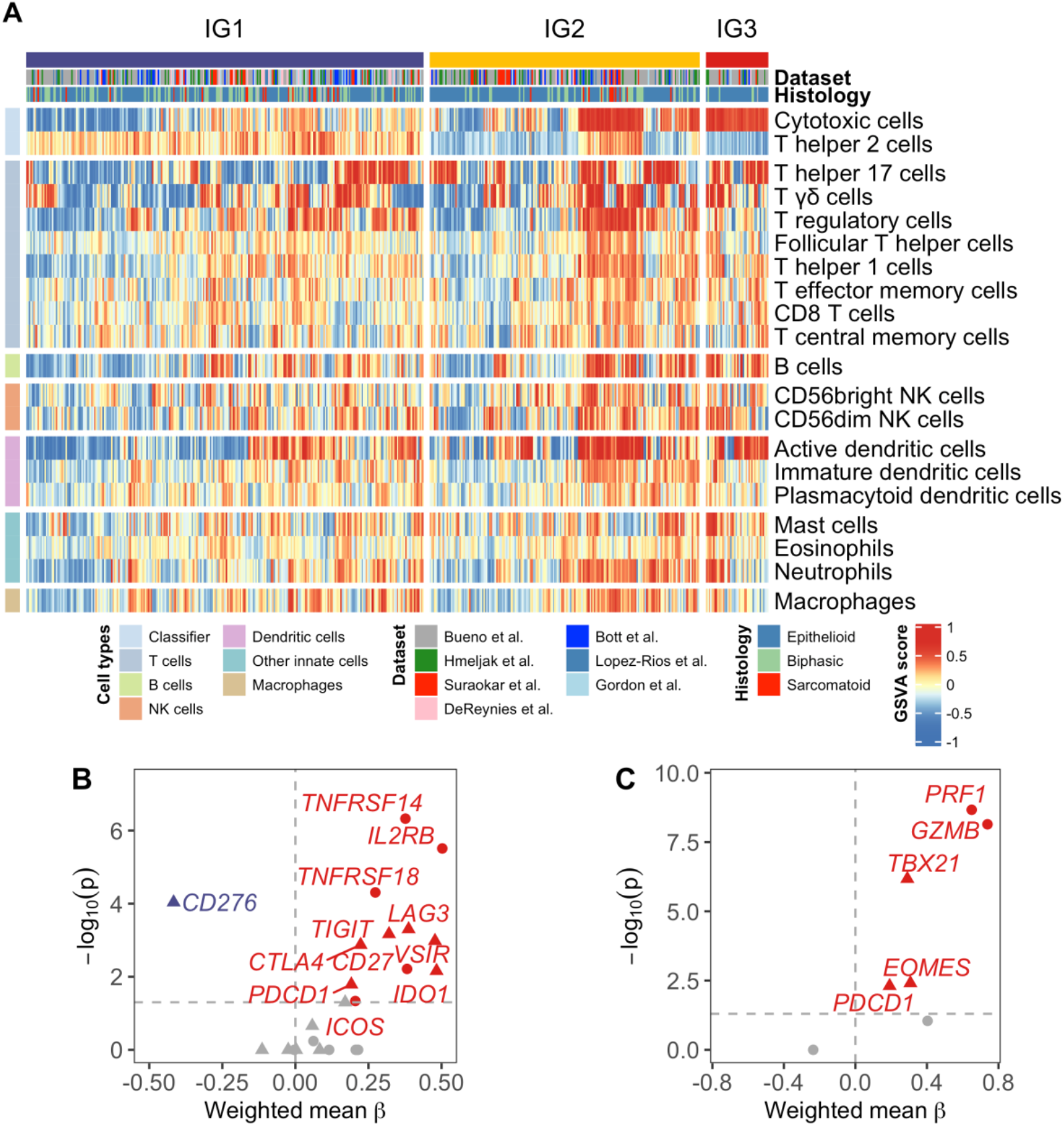
A) Relative abundances of the 20 immune fractions among 516 MPM tumours. Samples are stratified in each group and clustered using Ward’s method and Euclidean distance. B) Volcano plot of immune checkpoints inhibitors (triangle) and activators (dot). Effect size (β) correlates with immune groups: negative β values correspond to decreasing expression from IG1 to IG3, while positive values indicate increasing expression from IG1 to IG3. Grey-coloured dots are not significant. C) Volcano plot of T-cell exhaustion (triangle) and effector (dot) markers. Effect size (β) correlates with immune groups. Grey-coloured dots are not significant.

In order to explore potential additional clinical correlates of the three identified MPM immune groups, we analyzed their association with available clinical covariates: age, sex, tumor stage at the time of diagnosis, tumor histology, asbestos exposure, and neoadjuvant therapy. Out of all the variables explored, we identified a significant association with age and histology (Table 1). Patients’ median age in group IG1 is 66 years, compared to 63.7 years and 63.2 years for groups IG2 and IG3, respectively (p=0.021). Regarding tumor histology, IG3 showed a strong enrichment in epithelioid tumors (90.70% in IG3 vs 62.04% in IG1 and 73.68% in IG2, p=0.001).

**Table 1.** Summary of clinicopathological variables by immune group.

|  | N | IG1<br>N=281 | IG2<br>N=191 | IG3<br>N=44 | p-value |
| --- | --- | --- | --- | --- | --- |
| Age, Median (Range) | 385 | 66.00<br>(28.00 - 86.00) | 63.70<br>(30.20-84.50) | 63.20<br>(18.80-76.00) | 0.021 |
| Sex, N (%): | 385 |  |  |  | 0.115 |
| Male |  | 166 (83.42%) | 120 (81.63%) | 27 (69.23%) |  |
| Female |  | 33 (16.58%) | 27 (18.37%) | 12 (30.77%) |  |
| Stage, N (%): | 348 |  |  |  | 0.888 |
| Stage I |  | 10 (5.32%) | 9 (7.26%) | 3 (8.33%) |  |
| Stage II |  | 33 (17.55%) | 21 (16.94%) | 7 (19.44%) |  |
| Stage III |  | 108 (57.45%) | 68 (54.84%) | 22 (61.11%) |  |
| Stage IV |  | 37 (19.68%) | 26 (20.97%) | 4 (11.11%) |  |
| Histology, N (%): | 507 |  |  |  | 0.001 |
| Epithelioid |  | 170 (62.04%) | 140 (73.68%) | 39 (90.70%) |  |
| Biphasic |  | 84 (30.66%) | 39 (20.53%) | 4 (9.30%) |  |
| Sarcomatoid |  | 20 (7.30%) | 11 (5.79%) | 0 (0.00%) |  |
| Asbestos exposure, N (%): | 351 |  |  |  | 0.068 |
| No |  | 41 (22.53%) | 42 (31.82%) | 15 (40.54%) |  |
| Possible |  | 1 (0.55%) | 2 (1.52%) | 1 (2.70%) |  |
| Yes |  | 140 (76.92%) | 88 (66.67%) | 21 (56.76%) |  |
| Neoadjuvant therapy, N (%): | 325 |  |  |  | 0.11 |
| No |  | 163 (93.68%) | 101 (87.07%) | 30 (85.71%) |  |
| Yes |  | 11 (6.32%) | 15 (12.93%) | 5 (14.29%) |  |

To further exclude a potential confounding role of the tumor histology on the differential survival among the immune groups, we evaluated whether the immune-based classification still was associated with patient survival within both epithelioid and non-epithelioid tumors. In epithelioid tumors, we observed improved survival in I_G2_ and I_G3_ patients compared to I_G1_ (HR_IG2_=0.47, HR_IG3_=0.24, p=3.310^-8^) (Suppl. Fig. S3A). For non-epithelioid tumors, the very small number of IG3 patients (n=3) prevented us from obtaining reliable results for this subgroup, but we still observed a trend towards better survival in IG2 patients compared to IG1 (HR_IG2_=0.60, p=0.074) (Suppl. Fig. S3B).

### Immune groups correlate with most immune checkpoint markers and markers of effector and exhausted T cells

To further evaluate the potential clinical relevance of the identified immune groups, we evaluated the expression of a wide selection of immune checkpoint inhibitors and activators. We observed a significant increasing expression pattern across MPM immune groups for most of the evaluated checkpoint markers (Fig. 2B). *TNFRSF14, IL2RB, TNFRSF18 (GITR), CD27* and *ICOS* were among the strongest positively associated activator markers with the immune groups. Regarding immune checkpoint inhibitors, *LAG3, TIGIT, VSIR (VISTA), IDO1, CTLA4* and *PDCD1 (PD-1)* were the strongest correlated markers. In opposition, *CD276 (B7-H3)* showed negative correlation with the immune groups, being its expression higher in IG1 tumors than tumors in IG2 and IG3 groups. Detailed expression of *CD274 (PD-L1), PDCD1 (PD-1), CTLA4, CD276 (B7-H3)*, and *VSIR (VISTA)* in each dataset is available in Suppl. Fig. S4A, showing the distribution of tumors in each group. An overall higher expression of *VSIR* with respect to the other immune checkpoints is observed, as previously reported in other publications. [7,32] Importantly, immune groups still show significant differences in prognosis independently of the expression level of any of these five immune checkpoints (Suppl. Fig. S4B). Additionally, we also evaluated markers of effector and exhausted T cells correlated with the identified MPM immune groups (Fig. 2C). In our analysis, all evaluated markers - excluding *CD44* - showed a positive correlation with the immune group classification, with a majority being significantly enriched in IG3 *(PRF1, GZB, TBX21, EOMES, PDCD1)*.

### Genomic characterization reveals IG1 tumors present more genomic instability

Once we identified the different immune-based groups in MPM tumors and established their potential clinical relevance, we wanted to characterize the different groups at the genomic level to identify any potential differential genomic patterns. These analyses were performed using Bueno et al.[6] and Hmeljak et al.[7] datasets. Using whole-exome data from Bueno et al.[6] and Hmeljak et al.[7] datasets, we assessed if immune groups were different in terms of TMB, mutational signatures,[33] or antigen immunogenicity (Fig. 3A-C). Although we did not find statistically significant differences among immune groups for any of the previous analyses, we did observe a slight decrease of mutational Signature 3 in IG3 tumors compared to IG1 and IG2 patients. Signature 3 is associated with homologous recombination deficiency and *BRCA1/2* deficiency.[33] Additionally, at the singlegene level no gene was found to be preferentially mutated in any group (data not shown).

**Figure 3.**
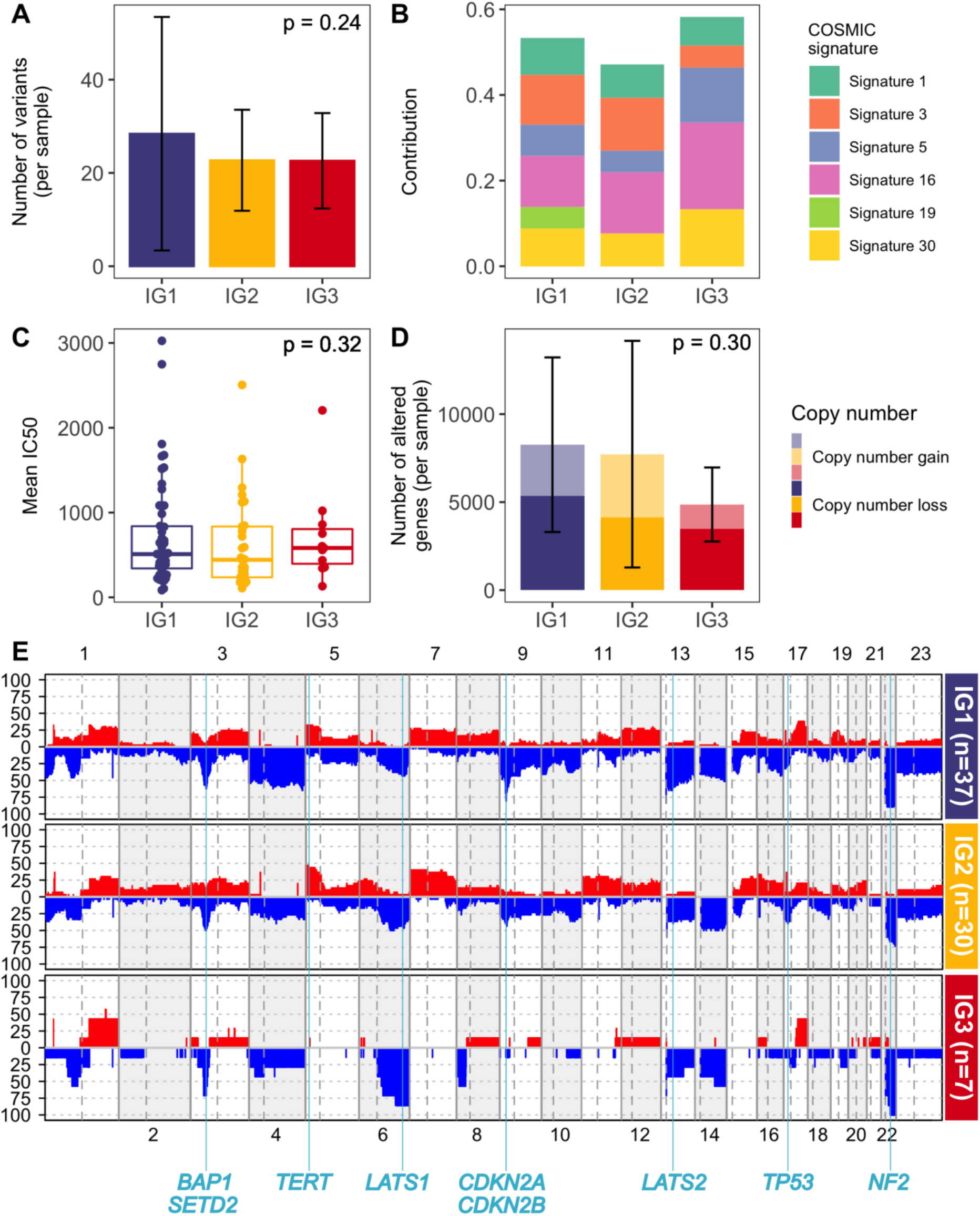
Genomic characterization A) Tumour mutational burden among immune groups. Mean number of variants per sample. Error bars indicate SD. B) Distribution of COSMIC mutational signatures among each immune group. C) Immunogenicity as the mean of IC50 affinity of mutations among immune groups. D) Copy number burden among immune groups. Mean number of altered genes per sample. Error bars indicate SD. Lighter colours show copy number gains while darker ones depict copy number losses. E) Genomic overview showing the percentage of samples with copy number alterations among each immune group. Landmark MPM genes are depicted according to their genomic location.

Copy number alterations were assessed for Hmeljak et al. dataset.[7] There were no significant differences between immune groups in the overall number of altered genes per sample, although tumors in IG3 tend to have fewer copy number alterations overall compared to the other groups as shown in Figure 3D. Figure 3E shows the overall percentage of altered samples across the genome for each immune group. Regarding MPM landmark genes, genes like *BAP1, SETD2, LATS2, TP53* and *NF2* did not show differences between groups. *CDKN2A* however is altered in 81% of tumors in IG1, in 47% of tumors in IG2, and 14% of tumors in IG3. Suppl. Fig. S5 shows expression of frequent tumor suppressor genes inactivated in MPM,[3] and results correlate with genomics, as *CDKN2A* expression is strongly correlated with immune groups in the largest datasets.

### Transcriptional cell cycle / immune system activity trade-off across immune groups

Next, we sought for gene expression differences among the three immune groups, to identify those genes with an increasing or decreasing pattern among the three groups. The results are depicted in Fig. 4A, which shows a high number of genes with decreasing expression from IG1 to IG3 tumors related to cell proliferation such as *CENPF* (centromere protein also known as mitosin), *BIRC5* (otherwise known as survivin, preventing apoptotic cell death), or kinesins like *KIF23*, involved in cell division. On the other hand, genes positively correlated with the immune groups include immune system related genes such as *HSH2D*, a target of T-cell activation signaling pathways, and *GNLY*, which encodes for a protein present in cytotoxic granules of cytotoxic T lymphocytes.

**Figure 4.**
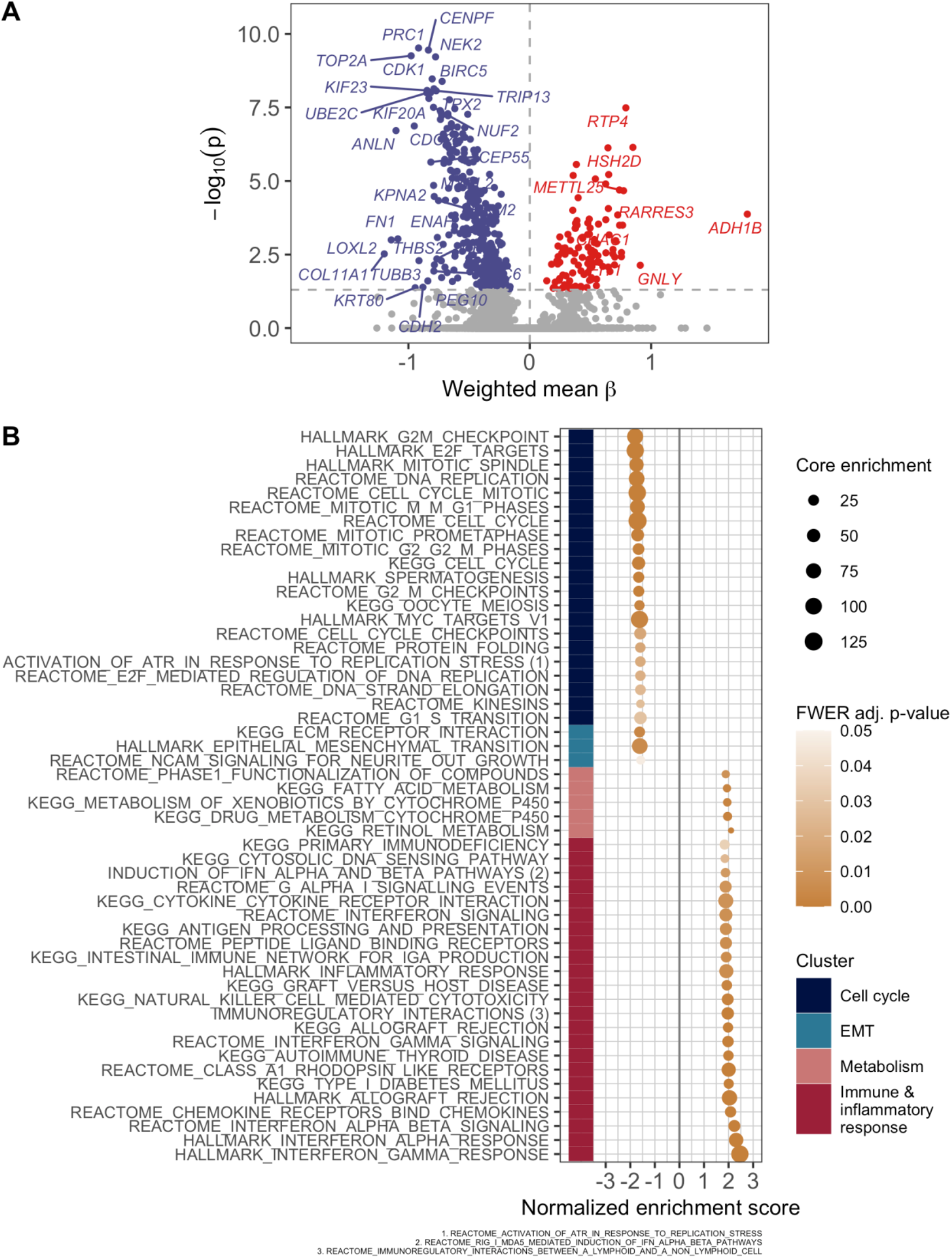
Transcriptomic characterization A) Volcano plot of gene expression. Effect size (β) correlates with immune groups. B) Significant pathways from pre-ranked GSEA. Positive normalized enrichment scores correlate with an increasing expression of pathway from IG1 to IG3, while negative ones depict higher pathway expression for IG1 tumours vs. IG3 tumours. Pathways are clustered using Ward’s method and an odds-ratio based distance.

In order to capture more precisely the enriched pathways and biological functions of the genes identified in the previous analysis, we performed a pre-ranked GSEA using a set of transcriptional signatures covering a wide range of pathways, and the genes ranked according to their linear association with the immune groups. The results are shown in Fig. 4B and we observed that signatures more positively correlated with the immune group (i.e., more expressed in IG3 tumors) were mostly related to immune system and inflammatory response processes. Contrarily, signatures negatively correlated with the immune groups mostly relate to cell cycle and epithelial mesenchymal transition, suggesting a trade-off between these molecular processes and the immune system.

Finally, since IG1 tumors appeared to be driven by cell cycle deregulation, we assessed the potential confounder effect among cell proliferation and immune groups. Therefore, we tested the association between the cell proliferation marker *MKI67* and the immune groups. Notably, regardless of *MKI67* expression levels, the association of the improving survival pattern across immune groups is upheld (Suppl. Fig. S6).

### Potential therapeutic strategies

To further evaluate the potential clinical benefit of the immune-based MPM classification, we decided to assess the behavior of different treatment-response signatures among the three groups. We first wanted to identify whether there were any differences in the response to the currently available standard chemotherapy treatment based on the combination of cisplatin and pemetrexed. We obtained two expression-based signatures derived from non-small cell lung cancer that predicted resistance to pemetrexed[21] and to cisplatin.[22] Figure 5A displays GSVA scores for the two signatures across the three MPM immune groups. Overall, the results of the analysis suggested that MPM tumors in IG1 might be more resistant to pemetrexed, while IG3 tumors show a trend to lower resistance. On the other hand, cisplatin resistance analysis showed no differences between immune groups.

**Figure 5.**
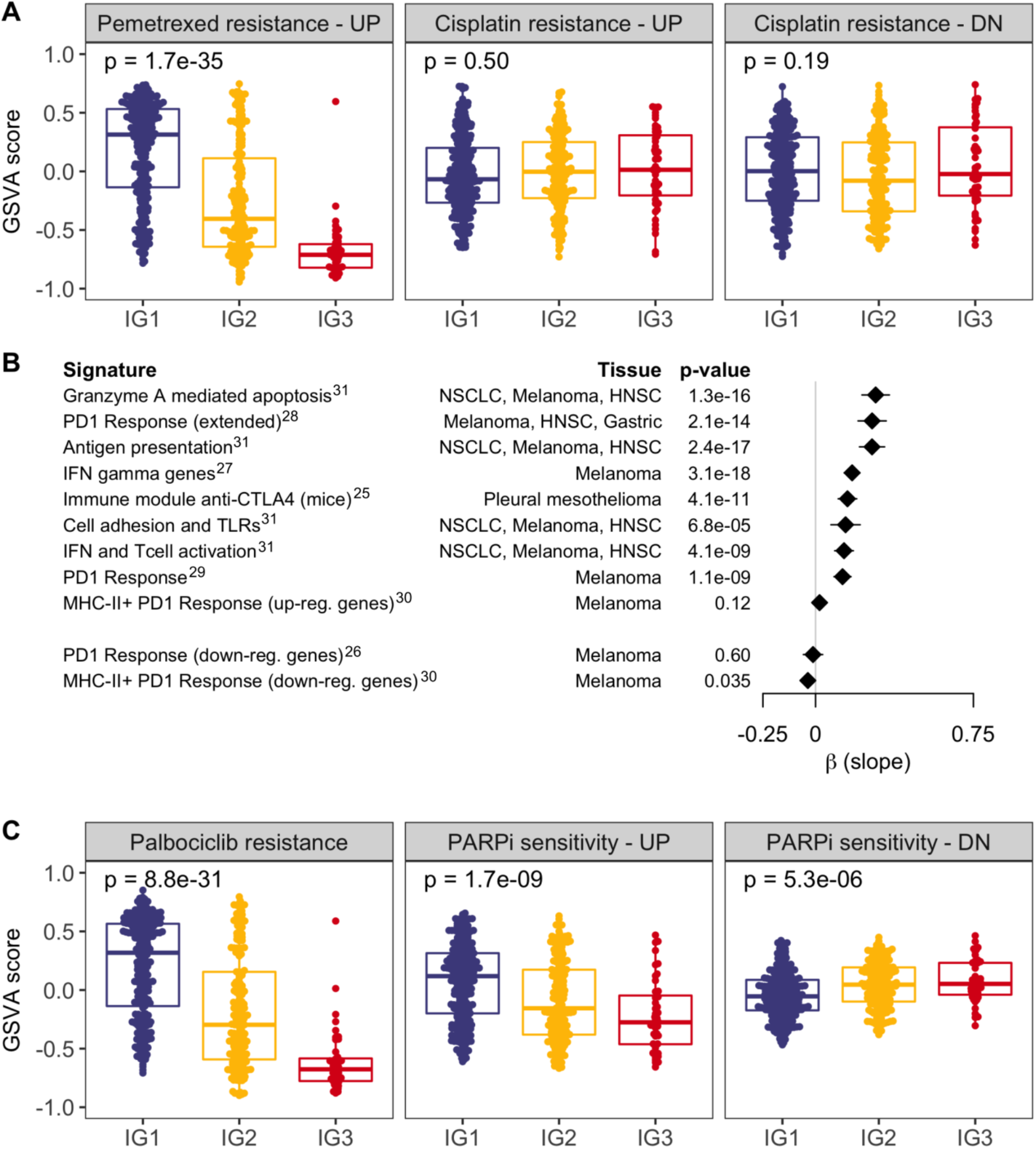
Assessment of gene expression signatures predictive of benefit or resistance to multiple treatments. GSVA scores for each sample allowed to test resistance to first-line chemotherapy (A); response to immunotherapy (B), and resistance or sensitivity to targeted therapy (C). Superscripts in immunotherapy signatures correspond to bibliographic references. Up-reg.: up-regulated; down-reg.: down-regulated.

Next, we tested a large set of gene-expression signatures predictive of benefit from ICI treatment[25-31] to formally evaluate if MPM tumors in IG3 could benefit from an immune-based therapeutic approach (Fig. 5B). The overlap between these signatures and the classifier is almost non-existent (3 shared genes at most). We observed that all signatures were positively correlated with the immune groups, with IG3 tumors presenting a significant upregulation for all but one of the signatures. Given the increased activation of cell cycle and DNA repair processes in the IG1 group of MPM tumors, we wanted to explore whether any differences in sensitivity to cyclin dependent kinase inhibitors and to drugs that inhibit DNA repair among the three groups. To do that, we collected a signature of resistance to palbociclib derived from breast cancer[24] and another computationally-derived signature predicting sensitivity to DNA repair inhibition with PARP inhibitors.[23] (Fig. 5C). Despite a higher cycling nature of tumors in group IG1, they appear to be more resistant to palbociclib than tumors in other groups, based on this predictive signature. Strikingly, tumors in IG1 showed increased sensitivity to PARP inhibitors compared to the other groups.

## DISCUSSION

In this study we performed a comprehensive analysis of publicly available transcriptome data from more than 500 MPM tumors in order to characterize the immune landscape of this disease. By the identification of immune cells associated with clinical outcome, we classified MPM tumors according to their T-helper 2 and cytotoxic T cell abundance levels. This classification stratifies patients into three groups that represent different immune infiltration patterns and are associated with distinct survival outcomes. The group with the shortest overall survival, IG1, represents more than 50% of the analyzed tumors, while IG3, with better prognosis, accounts for only 8.5% of the tumors. We observed an increasing pattern of abundance for most immune fractions across the three immune groups. Interestingly, the survival benefit of this classification is consistent in both epithelioid and non-epithelioid tumors. We further characterized these three MPM immune groups at the genomic and transcriptomic levels, and identified potential therapeutic strategies using predictive signatures and large-scale pharmacogenomics data.

Different immune-related MPM phenotypes have already been described in recent studies using comprehensive approaches.[34-38] These studies use different techniques such as Nanostring™ technology,[35] mass cytometry,[34] or RNA-sequencing.[37] The diversity of obtained results suggests that the composition and role of the tumor microenvironment in MPM is remarkably complex and controversial, and thus further studies will contribute to elucidate this question.

In our work, MPM tumors are classified based on T_H2_ and T_C_ cells abundance, as previously stated. T_H2_ are CD4+ T cells which are induced by the presence of interleukin 4 (IL4) via STAT6 signaling and regulate humoral immune responses and responses to extracellular pathogens. T_H2_ cells secrete IL4, IL5, IL10 and IL13 which promote immune suppression by inhibiting T-Helper 1 cytokine production. Additionally, a recent study identified higher levels of IL5 and IL13 after exposure to asbestos in mesothelial cells.[39] The role of T_H2_ cells in cancer has been found to be dual and context-dependent. While they can generate antitumor immunity by recruiting specific populations of innate immune cells, they have also been found to increase tumorigenicity in certain experimental models.[40] Consistent with our results, higher levels of T_H2_ cells have already been associated with poor prognosis in multiple cancer types.[41-43] Finally, in the analysis performed by Hmeljak et al.,[7] this signature has already been associated to the group with the worst prognosis.

On the other hand, CD8+ T_C_ cells are essential actors of the effector function of adaptive cellular immune response.[44] Along with natural killer (NK) cells, T_C_ cells are ultimately responsible for targeting and attacking cancer cells by secreting cytotoxins (e.g., granzymes and perforins) which reach the tumor cell cytoplasm and trigger a caspase-mediated apoptotic process.[45] Additionally, in agreement with our study, a recent pan-cancer study observed that patients with tumors with higher T_C_ infiltration tended to have better survival than patients with less Tc-infiltrated tumors.[46]

While most immune checkpoints correlate with the defined immune groups, *CD276 (B7-H3)* shows an opposite pattern of expression, with decreasing expression from IG1 to IG3. *CD276* is a member of the B7 family of immunoregulatory proteins and is overexpressed in distinct tumor types. It has been shown that *CD276* can promote tumor proliferation, angiogenesis and metastasis and is associated with shorter survival time.[47] This result ties in with the survival pattern among the groups as well as with results from functional analysis in which cell cycle shows a decreasing pattern from IG1 to IG3. Similarly, *CD44* is the only T-cell exhaustion marker that shows negative correlation with the immune groups. This marker has been associated with metastasis and low survival rate in multiple cancer types, and chemoresistance in prostate and head and neck squamous cell cancer types.[48] In MPM, *CD44* has been shown to promote invasiveness when interacting with hyaluronan.[49]

Regarding molecular characteristics, immune groups were not significantly correlated with specific genomic alterations in terms of gene mutations or copy number alterations, yet there seems to be a pattern of decreasing genomic instability among the groups. Genomic instability, higher in IG1, is also supported by the analysis of mutational signatures, revealing a possible stronger presence of DNA damage repair deficiency by means of homologous recombination repair. In terms of copy number, no global differences were found among MPM groups, possibly due to the smaller sample size (only one dataset with available data). However, genomic regions like 9p21, harboring landmark genes in MPM as *CDKN2A* or type I interferon gene cluster, were likely to be lost in the IG1 compared to the other two groups. Transcriptomic characterization of these groups by gene enrichment analysis revealed a trade-off between immune response activity and cell proliferation mechanisms, showing higher activation of cell cycle in IG1 tumors, in agreement with genomic characteristics. This trade-off has already been described in other studies, reinforcing the hypothesis that cell cycle regulators and genomic instability could also have an impact on immune checkpoints.[46,50]

Finally, the lack of effective therapeutic strategies beyond chemotherapy in MPM calls for assessment of potential options and tailoring for each subset of patients. Currently several phase III clinical trials are assessing the role of ICI in combination with chemotherapy in the first line setting of patients with advanced MPM. The results of the CheckMate-743 trial evaluating nivolumab plus ipilimumab versus platinum-based chemotherapy in previously untreated MPM are eagerly awaited, since this trial met its primary endpoint of overall survival. In this sense, the treatment landscape of advanced MPM is likely to evolve and predictive markers of ICI benefit are needed. In our work, we observed that patients belonging to IG3 may benefit from pemetrexed and immunotherapy, while no group showed any strong predisposition to benefit from chemotherapy with cisplatin treatment. Moreover, tumors in IG1 appear to be more sensitive to PARP inhibitors. This coincides with a higher proportion of genomic instability and mutational signature 3, associated with BRCA deficiency. It is important to stress that this classification needs further validation in a prospective cohort to identify the most suitable therapeutic strategy for patients in each immune group. It is also important to point out that this study focused on gene expression signatures as proxies since there is no information regarding immunotherapy in large pharmaco-genomic assays and also that even though pemetrexed and cisplatin are commonly administered in combination; we assessed their effect as single therapies due to the lack of signatures for the combined treatment.

To sum up, this study identifies a novel signature with potential clinical relevance based on TH2 and Tc levels and unveils a small fraction of MPM tumors that show better prognosis. These groups have different characteristics, both at the genomic and transcriptomic levels, and these differences could be used to tailor potential therapies for each group in the future. Further research is needed towards the identification of a reduced signature valid for FFPE samples, and validation of these results is warranted in an independent and prospective cohort of MPM tumors preferentially treated with ICI.

## Data Availability

Data used in this study are available in public, open access repositories.

## Declarations

## Acknowledgements

We thank CERCA Programme / Generalitat de Catalunya for institutional support.

We thank all the researchers who kindly put their research data for public use. Bueno et al. data was accessed through a data transfer agreement with Genentech (DAT-015429).

## Funding

This work is supported by the Carlos III National Health Institute funded by FEDER funds - a way to build Europe - [PI14/01109; PI18/00920]; the Government of Catalonia [2017SGR448].

XS is supported by RTI2018-102134-A-I00 grant funded by Spanish Ministry of Science and Innovation. EN received support from the SLT006/17/00127 grant, funded by the Department of Health of the Generalitat de Catalunya by the call “Acció instrumental d’intensificació de professionals de la salut”. This study has been funded by Sociedad Española de Oncología Médica (SEOM) through “Proyectos de Investigación para Grupo Emergente”.

## Authors’ contributions

XS and EN conceived and designed the study. AA and DC performed the computational and statistical analysis. XS, EN, AA, DC, SHP, and EA contributed to the interpretation of the results. XS, EN, AA, and DC drafted the manuscript. ALD, RM, RP, RL, IE, RR, SP, and VM provided critical revisions of the article. All authors read and approved the final manuscript.

## Competing interests

VM is consultant to Bioiberica S.A.U. and Grupo Ferrer S.A., received research funds from Universal DX, and is coinvestigator in grants with Aniling. EN participated in advisory boards from Bristol Myers Squibb, Merck Sharpe & Dohme, Lilly, Roche, Pfizer, Takeda, Boehringer Ingelheim, Amgen and AstraZeneca. The other authors have no conflicts of interest to declare.

## Ethics approval and consent to participate

Not required.

## Data availability statement

Data used in this study are available in public, open access repositories.

List of abbreviations
FDR: False Discovery Rate
GSEA: Gene Set Enrichment Analysis
GSVA: Gene Set Variation Analysis
ICI: Immune Checkpoint Inhibitors
MPM: Malignant Pleural Mesothelioma
PD1: Programmed Cell Death Protein 1
PD-L1: Programmed Death Ligand 1
T_C_: Cytotoxic T cells
T_H2_: T-helper 2 cells
TMB: Tumor Mutational Burden

## REFERENCES

1 Meyerhoff RR, Yang C-FJ, Speicher PJ, et al. Impact of mesothelioma histologic subtype on outcomes in the Surveillance, Epidemiology, and End Results database. J Surg Res 2015;196:23–32. doi:10.1016/j.jss.2015.01.043

2 Kindler HL, Ismaila N, Armato SG, et al. Treatment of Malignant Pleural Mesothelioma: American Society of Clinical Oncology Clinical Practice Guideline. J Clin Oncol 2018;36:1343–73. doi:10.1200/JC0.2017.76.6394

3 Yap TA, Aerts JG, Popat S, et al. Novel insights into mesothelioma biology and implications for therapy. Nat Rev Cancer 2017;17:475–88. doi:10.1038/nrc.2017.42

4 McCambridge AJ, Napolitano A, Mansfield AS, et al. Progress in the Management of Malignant Pleural Mesothelioma in 2017. J Thorac Oncol 2018;13:606–23. doi:10.1016/j.jtho.2018.02.021

5 Scherpereel A, Wallyn F, Albelda SM, et al. Novel therapies for malignant pleural mesothelioma. Lancet Oncol 2018;19:e161-72. doi:10.1016/S1470-2045(18)30100-1

6 Bueno R, Stawiski EW, Goldstein LD, et al. Comprehensive genomic analysis of malignant pleural mesothelioma identifies recurrent mutations, gene fusions and splicing alterations. Nat Genet 2016;48:407–16. doi:10.1038/ng.3520

7 Hmeljak J, Sanchez-Vega F, Hoadley KA, et al. Integrative Molecular Characterization of Malignant Pleural Mesothelioma. Cancer Discov 2018;8:1548–65. doi:10.1158/2159-8290.CD-18-0804

8 Suraokar MB, Nunez MI, Diao L, et al. Expression profiling stratifies mesothelioma tumors and signifies deregulation of spindle checkpoint pathway and microtubule network with therapeutic implications. Ann Oncol 2014;25:1184–92. doi:10.1093/annonc/mdu127

9 de Reyniès A, Jaurand M-C, Renier A, et al. Molecular classification of malignant pleural mesothelioma: identification of a poor prognosis subgroup linked to the epithelial-to-mesenchymal transition. Clin Cancer Res 2014;20:1323–34. doi:10.1158/1078-0432.CCR-13-2429

10 Bott M, Brevet M, Taylor BS, et al. The nuclear deubiquitinase BAP1 is commonly inactivated by somatic mutations and 3p21.1 losses in malignant pleural mesothelioma. Nat Genet 2011;43:668–72. doi:10.1038/ng.855

11 López-Ríos F, Chuai S, Flores R, et al. Global gene expression profiling of pleural mesotheliomas: overexpression of aurora kinases and P16/CDKN2A deletion as prognostic factors and critical evaluation of microarray-based prognostic prediction. Cancer Res 2006;66:2970–9. doi:10.1158/0008-5472.CAN-05-3907

12 Gordon GJ, Rockwell GN, Jensen RV, et al. Identification of novel candidate oncogenes and tumor suppressors in malignant pleural mesothelioma using large-scale transcriptional profiling. Am J Pathol 2005;166:1827–40. doi:10.1016/S0002-9440(10)62492-3

13 Van der Auwera GA, Carneiro MO, Hartl C, et al. From FastQ data to high confidence variant calls: the Genome Analysis Toolkit best practices pipeline. Curr Protoc Bioinformatics 2013;43:11.10.1-11.10.33. doi:10.1002/0471250953.bi1110s43

14 Irizarry RA, Hobbs B, Collin F, et al. Exploration, normalization, and summaries of high density oligonucleotide array probe level data. Biostatistics 2003;4:249–64. doi:10.1093/biostatistics/4.2.249

15 Hänzelmann S, Castelo R, Guinney J. GSVA: gene set variation analysis for microarray and RNA-seq data. BMC Bioinformatics 2013;14:7. doi:10.1186/1471-2105-14-7

16 Bindea G, Mlecnik B, Tosolini M, et al. Spatiotemporal dynamics of intratumoral immune cells reveal the immune landscape in human cancer. Immunity 2013;39:782–95. doi:10.1016/j.immuni.2013.10.003

17 Charoentong P, Finotello F, Angelova M, et al. Pan-cancer Immunogenomic Analyses Reveal Genotype-Immunophenotype Relationships and Predictors of Response to Checkpoint Blockade. Cell Rep 2017;18:248–62. doi:10.1016/j.celrep.2016.12.019

18 Pardoll DM. The blockade of immune checkpoints in cancer immunotherapy. Nat Rev Cancer 2012; 12:252-64. doi:10.1038/nrc3239

19 Wherry EJ, Kurachi M. Molecular and cellular insights into T cell exhaustion. Nat Rev Immunol 2015;15:486–99. doi:10.1038/nri3862

20 Subramanian A, Tamayo P, Mootha VK, et al. Gene set enrichment analysis: a knowledge-based approach for interpreting genome-wide expression profiles. Proc Natl Acad Sci USA 2005;102:15545–50. doi:10.1073/pnas.0506580102

21 Hou J, Lambers M, den Hamer B, et al. Expression profiling-based subtyping identifies novel non-small cell lung cancer subgroups and implicates putative resistance to pemetrexed therapy. J Thorac Oncol 2012;7:105–14. doi:10.1097/JT0.0b013e3182352a45

22 Whiteside MA, Chen D-T, Desmond RA, et al. A novel time-course cDNA microarray analysis method identifies genes associated with the development of cisplatin resistance. Oncogene 2004;23:744–52. doi:10.1038/sj.onc.1207164

23 McGrail DJ, Lin CC-J, Garnett J, et al. Improved prediction of PARP inhibitor response and identification of synergizing agents through use of a novel gene expression signature generation algorithm. NPJ Syst Biol Appl 2017;3:8. doi:10.1038/s41540-017-0011-6

24 Malorni L, Piazza S, Ciani Y, et al. A gene expression signature of retinoblastoma loss-of-function is a predictive biomarker of resistance to palbociclib in breast cancer cell lines and is prognostic in patients with ER positive early breast cancer. Oncotarget 2016;7:68012–22. doi:10.18632/oncotarget.12010

25 Lesterhuis WJ, Rinaldi C, Jones A, et al. Network analysis of immunotherapy-induced regressing tumours identifies novel synergistic drug combinations. Sci Rep 2015;5:12298. doi:10.1038/srep12298

26 Hugo W, Zaretsky JM, Sun L, et al. Genomic and Transcriptomic Features of Response to Anti-PD-1 Therapy in Metastatic Melanoma. Cell 2016;165:35–44. doi:10.1016/j.cell.2016.02.065

27 Gao J, Shi LZ, Zhao H, et al. Loss of IFN-γ Pathway Genes in Tumor Cells as a Mechanism of Resistance to Anti-CTLA-4 Therapy. Cell 2016;167:397–404.e9. doi:10.1016/j.cell.2016.08.069

28 Ayers M, Lunceford J, Nebozhyn M, et al. IFN-γ-related mRNA profile predicts clinical response to PD-1 blockade. J Clin Invest 2017;127:2930–40. doi:10.1172/JCI91190

29 Chen P-L, Roh W, Reuben A, et al. Analysis of Immune Signatures in Longitudinal Tumor Samples Yields Insight into Biomarkers of Response and Mechanisms of Resistance to Immune Checkpoint Blockade. Cancer Discov 2016;6:827–37. doi:10.1158/2159-8290.CD-15-1545

30 Johnson DB, Estrada MV, Salgado R, et al. Melanoma-specific MHC-II expression represents a tumour-autonomous phenotype and predicts response to anti-PD-1/PD-L1 therapy. Nat Commun 2016;7:10582. doi:10.1038/ncomms10582

31 Prat A, Navarro A, Paré L, et al. Immune-Related Gene Expression Profiling After PD-1 Blockade in Non-Small Cell Lung Carcinoma, Head and Neck Squamous Cell Carcinoma, and Melanoma. Cancer Res 2017;77:3540–50. doi:10.1158/0008-5472.CAN-16-3556

32 Muller S, Victoria Lai W, Adusumilli PS, et al. V-domain Ig-containing suppressor of T-cell activation (VISTA), a potentially targetable immune checkpoint molecule, is highly expressed in epithelioid malignant pleural mesothelioma. Mod Pathol 2020;33:303–11. doi:10.1038/s41379-019-0364-z

33 Alexandrov LB, Nik-Zainal S, Wedge DC, et al. Signatures of mutational processes in human cancer. Nature 2013;500:415–21. doi:10.1038/nature12477

34 Lee H-S, Jang H-J, Choi JM, et al. Comprehensive immunoproteogenomic analyses of malignant pleural mesothelioma. JCI Insight 2018;3. doi:10.1172/jci.insight.98575

35 Patil NS, Righi L, Koeppen H, et al. Molecular and Histopathological Characterization of the Tumor Immune Microenvironment in Advanced Stage of Malignant Pleural Mesothelioma. J Thorac Oncol 2018;13:124–33. doi:10.1016/j.jtho.2017.09.1968

36 Blum Y, Meiller C, Quetel L, et al. Dissecting heterogeneity in malignant pleural mesothelioma through histo-molecular gradients for clinical applications. Nat Commun 2019;10:1333. doi:10.1038/s41467-019-09307-6

37 Alcala N, Mangiante L, Le-Stang N, et al. Redefining malignant pleural mesothelioma types as a continuum uncovers immune-vascular interactions. EBioMedicine 2019;48:191–202. doi:10.1016/j.ebiom.2019.09.003

38 Sneddon S, Rive CM, Ma S, et al. Identification of a CD8+ T-cell response to a predicted neoantigen in malignant mesothelioma. OncoImmunology 2020;9:1684713. doi:10.1080/2162402X.2019.1684713

39 Maki Y, Nishimura Y, Toyooka S, et al. The proliferative effects of asbestos-exposed peripheral blood mononuclear cells on mesothelial cells. Oncol Lett 2016;11:3308–16. doi:10.3892/ol.2016.4412

40 Liudahl SM, Coussens LM. To Help or To Harm. In: Immunology. Elsevier 2018. 97-116. doi:10.1016/B978-0-12-809819-6.00008-3

41 De Monte L, Reni M, Tassi E, et al. Intratumor T helper type 2 cell infiltrate correlates with cancer-associated fibroblast thymic stromal lymphopoietin production and reduced survival in pancreatic cancer. J Exp Med 2011;208:469–78. doi:10.1084/jem.20101876

42 Kusuda T, Shigemasa K, Arihiro K, et al. Relative expression levels of Th1 and Th2 cytokine mRNA are independent prognostic factors in patients with ovarian cancer. Oncol Rep 2005;13:1153–8.

43 Nevala WK, Vachon CM, Leontovich AA, et al. Evidence of systemic T_H2_-driven chronic inflammation in patients with metastatic melanoma. Clin Cancer Res 2009;15:1931–9. doi:10.1158/1078-0432.CCR-08-1980

44 Andersen MH, Schrama D, Thor Straten P, et al. Cytotoxic T cells. J Invest Dermatol 2006;126:32–41. doi:10.1038/sj.jid.5700001

45 Martínez-Lostao L, Anel A, Pardo J. How Do Cytotoxic Lymphocytes Kill Cancer Cells? Clin Cancer Res 2015;21:5047–56. doi:10.1158/1078-0432.CCR-15-0685

46 Tamborero D, Rubio-Perez C, Muiños F, et al. A Pan-cancer Landscape of Interactions between Solid Tumors and Infiltrating Immune Cell Populations. Clin Cancer Res 2018;24:3717–28. doi:10.1158/1078-0432.CCR-17-3509

47 Seaman S, Zhu Z, Saha S, et al. Eradication of Tumors through Simultaneous Ablation of CD276/B7-H3-Positive Tumor Cells and Tumor Vasculature. Cancer Cell 2017;31:501–515.e8. doi:10.1016/j.ccell.2017.03.005

48 Chen C, Zhao S, Karnad A, et al. The biology and role of CD44 in cancer progression: therapeutic implications. J Hematol Oncol 2018;11:64. doi:10.1186/s13045-018-0605-5

49 Cortes-Dericks L, Schmid RA. CD44 and its ligand hyaluronan as potential biomarkers in malignant pleural mesothelioma: evidence and perspectives. Respir Res 2017;18:58. doi:10.1186/s12931-017-0546-5

50 Wellenstein MD, de Visser KE. Cancer-Cell-Intrinsic Mechanisms Shaping the Tumor Immune Landscape. Immunity 2018;48:399–416. doi:10.1016/j.immuni.2018.03.004

